# Open Science Practices: A cross-sectional, observational, and descriptive study in a Portuguese nursing higher education institution

**DOI:** 10.1101/2025.03.26.25324671

**Authors:** Ana Sabrina Sousa, Mafalda Lopes, Diana Rodrigues, Palmira Oliveira, Regina Pires, Sara Pinto, Pedro Melo, João Frias Rosa, Amparo Alves, Rosa Silva

**Author notes:** Corresponding author (AS).

## Abstract

Open Science has emerged as an epistemological paradigm that promotes open and dynamic access to knowledge, fostering the sharing of scientific findings, collaboration between academia and society, and enhancing innovation and transparency. However, comprehensive studies on the adoption of Open Science practices remain limited, particularly in the nursing academic context. This study aimed to identify the current Open Science practices within a higher nursing education institution community. A cross-sectional, observational, and descriptive study was conducted in two phases. In the first phase, a questionnaire was adapted and piloted through a focus group. In the second phase, the final version of the questionnaire, with 34 questions, was distributed digitally to 747 teachers, researchers and students.

The study included 35 respondents, predominantly faculty members, with 50% holding a Ph.D. and 61.1% affiliated with research groups. While 77.8% reported engaging with Open Science publications, fewer respondents demonstrated familiarity with practices such as study preregistration and open-source coding. The principles of Open Science were widely accepted, particularly those emphasizing ethics, the democratization of knowledge, and scientific collaboration. Participants frequently used tools like EbscoHost, Medline, and Scopus, whereas platforms such as Zenodo and OpenUP Hub remained underutilized. Participants highlighted training priorities, especially in open access publishing, data management, and the implementation of Open Science recommendations. A notable finding of this study was the low participation in the study, suggesting the need to enhance awareness among the higher education community regarding the importance of Open Science. Although all participants agreed with the principles of Open Science, most reported not knowing its policies and not all were familiar with the various formats.

This study emphasized the need for investment in skills training for Open Science practices and for in-depth studies on motivation, adherence and enabling conditions, particularly in the context of Higher Education.

## Introduction

The emergence of Open Science (OS) as an epistemological paradigm reflects a shift towards open and dynamic access to information, allowing researchers to share knowledge in a publicly accessible environment [1,2]. Within this framework, OS serves as a tool for knowledge sharing between the scientific community and society, ensuring that research aligns with user needs and adheres to the principles of findability, accessibility, interoperability, and reusability (FAIR), thus enhancing innovation and maximizing the social and economic impact of scientific knowledge [2–5].

Consequently, the OS ecosystem has undergone exponential growth, reflecting not only its increasing adoption and recognition by various research stakeholders, but also its role in enhancing research quality, transparency, knowledge translation, and socioeconomic impact [6,7]. Moreover, OS enables researchers to move beyond restricted and closed frameworks, facilitating broader dissemination of scientific work while ensuring greater rigor and transparency in research processes [1,6]. As a result, OS fosters open practices and processes for the creation, sharing, and utilization of scientific knowledge by research and development institutions, among others, aligning with the principles of “Open Science” and the “Right to Science,” namely concerning access and participation [4].

The ongoing evolution of this concept and its associated domains strengthens and advances accessibility to research findings, data, and methodologies, challenging traditional paradigms of scientific knowledge production and dissemination in an era increasingly driven by co-creation and digital transformation [4,8].

In higher education, OS represents a significant transformation in knowledge management, redefining the methodologies for the production, dissemination, and implementation of scientific knowledge [8,9]. This approach carries substantial implications, challenges, and opportunities for students, faculty, knowledge managers, institutional leaders, and policymakers. The increasing availability of open resources, coupled with the potential for global collaboration, is profoundly reshaping the landscape of scientific research [7,8].

Access to innovative platforms and tools enhances the management and dissemination of research data, promoting more open and transparent practices [2,9]. Engagement in open research networks facilitates interdisciplinary synergies and the development of large-scale collaborative projects, cultivating a culture of co-creation and excellence in scientific production [6,10]. Additionally, OS reinforces the third mission of universities and higher education institutions by ensuring public accessibility to research findings and promoting social responsibility [11,12].

OS is based on fundamental principles, including transparency in practices, methodologies, observations, and data collection; public availability, accessibility, and reuse of research outputs such as publications and datasets; transparency in scientific communication; and the utilization of web-based tools to facilitate scientific collaboration [3,13,14]. Key components of OS include open access, which ensures online availability and unrestricted access to peer-reviewed scientific results; open data, which involves the publication and reuse of research datasets; open code, which makes software accessible through open source-code licenses; open replicable research, which supports the independent replication of scientific findings; and OS networks and citizen science initiatives [3,15,16]. Additionally, the Organization for Economic Cooperation and Development (OECD) [17] underscores the benefits of OS, such as reduced costs in data creation and transfer, the reuse of data to stimulate further research, strengthened partnerships within research teams, improved quality and transparency in validation processes, accelerated knowledge transfer, and active citizen engagement in science.

Ultimately, OS constitutes a fundamental paradigm shift in research, profoundly shaping the practices of higher education institutions and research and development units, with profound and enduring implications. Historically, the production of scientific knowledge has been highly closed and fragmented. Within the OS framework, fostering a culture of civic empowerment and social responsibility is essential to bridge the gap between academia and society, thereby promoting closer collaboration among researchers, educators, and the broader public [18].

Given its growing importance, higher education institutions are encouraged to implement proactive strategies to advance OS and maximize its benefits [19]. However, the existing body of scientific evidence on this topic remains limited. There is a lack of comprehensive studies that map current OS practices within the academic community, including diagnostic analyses of these practices, existing levels of knowledge, and the perceived needs of stakeholders in this domain, particularly in the nursing field. Thus, this study aimed to identify the prevailing OS practices within a higher education institution (HEI) community and, consequently, to assess its training and infrastructure requirements. The findings of this research may further contribute to the systematization of practices and the development of guidelines to enhance the implementation of OS best practices.

## Methodology

To ensure transparency and reproducibility, this study adhered to the guidelines established by the Strengthening the Reporting of Observational Studies in Epidemiology (STROBE) [20] throughout all stages, from study design to data analysis.

### Study Design

An observational, descriptive, and cross-sectional study was conducted at a public Portuguese Nursing HEI. The study design was structured in two main phases, which are presented below.

### Questionnaire Adaptation

The adapted questionnaire was derived from the Marques *et al.* [21] questionnaire and the Science Europe Principles on Open Access to Research Publications [22]. The adaptation process was led by a team member with recognized expertise in the field under study (ML). Following the adaptation process, the questionnaire was reviewed by two external experts, who suggested minor adjustments (FF, JA).

Subsequently, the questionnaire was piloted through a focus group with 10 participants in January 2024, including postgraduate students and faculty members. These participants were selected through convenience sampling from the research team’s network of contacts. This phase aimed at evaluating the content validity, appropriateness, and applicability of the questionnaire.

Three members of the research team participated in the focus group sessions, documenting all relevant information. The focus group reviewed the entire questionnaire, and changes were implemented based on the majority consensus. In instances where a definitive conclusion could not be reached, the information discussed during the sessions was carefully analyzed by the research team, and additional adjustments were made based on a consensus-driven approach.

### Questionnaire Application

The final version of the questionnaire was made available on a digital platform and distributed to the target population, which included faculty members and postgraduate students at the HEI, excluding undergraduate students (https://forms.office.com/e/qrGdPRkX36?origin=lprLink). The questionnaire comprised 34 questions with multiple-choice items and open-ended fields for free-text answers.

The data were gathered from 1^st^ February to 31^st^ May 2024. The sample represented an approximate population of 747 individuals (members at the institution allocate 30% of their teaching workload to research activities, except for adjunct lecturers). The student body consisted of healthcare professionals, specifically nurses enrolled in postgraduate or master’s programs. The questionnaire was disseminated through the institution’s internal communication channels, including email and newsletters.

### Data Collection Instrument

The variables examined in this study were organized into four main domains. The first domain, **Research Participation**, included aspects such as participation in research groups or centers and the participants’ roles in research projects. The second domain, **OS Practices**, focused on indicators like knowledge of OS, participation in OS projects, familiarity with open access policies, and practical experience with OS components, such as publications, educational materials, and data sharing. The third domain, **Tools and Platforms**, covered the use of bibliographic databases, institutional repositories, and specialized OS tools. Finally, the fourth domain, **Perceptions and Support Needs**, addressed variables such as the perceived advantages and disadvantages of OS, the main needs for support, and the sources of OS-related support utilized by participants. This information is presented in Table 1.

**Table 1.** Indicators Assessed in the Open Science Practices Questionnaire.

| Domain | Data collected |
| --- | --- |
| Participant characterization | Participation in Research Groups or Centers<br>Role in Research Projects |
| OS Practices | Knowledge of OS<br>Participation in OS Projects<br>Familiarity with Open Access Policies<br>Practical Experience with OS Components |
| Tools and Platforms | Use of Bibliographic Databases<br>Use of Institutional Repositories<br>Use of Specialized OS Tools |
| Perceptions and Support Needs | Perceived Advantages and Disadvantages<br>Main Needs for Support<br>Sources of OS Support |

### Ethical Procedures

The study was approved by the Nursing School of Porto ethics committee, ensuring full compliance with the General Data Protection Regulation (GDPR) to ensure anonymity (approval number CE_31/2023) in December 2023. Ethical approval was given by the board members: Elisabete Borges (president) and António Santos.

Participants were informed of the study’s objectives and provided written informed consent before completing the questionnaire. Identification codes were assigned to focus group participants to ensure anonymity and confidentiality (P1, P2, …). Data were securely stored on institutional servers, accessible only to the research team.

### Data Analysis

Information from the focus groups and open-ended questionnaire responses were submitted for qualitative analysis. The focus group data were transcribed and subjected to content analysis to identify emerging thematic categories, which were then discussed within the research team. Similarly, the open-ended responses were analyzed to identify relevant patterns and categories, which complemented the quantitative analysis.

Subsequently, a descriptive analysis of the variables was performed using IBM SPSS Statistics V.24 software. Incomplete responses were excluded to ensure the integrity and representativeness of the results.

## Results

### Questionnaire Adaptation

The focus group provided valuable insights to refine the questionnaire on OS practices. The modifications were categorized into three main areas: changes to questions, adjustments to the order and structure, and refinements in wording and response options, as detailed in Table 2. Key changes included the following:

**Table 2.** Categories Emerging from the Analysis.

| Category | Subcategory | Record unit |
| --- | --- | --- |
| Modifications to Questionnaire Questions | Addition of Questions | Add the question: "In your experience, what resources facilitate the adoption of Open Science practices?" with response options (Between questions 28 and 29). |
|  | Division of Questions | Split question 16: "We would like to ask you to identify up to 3 advantages and 3 disadvantages of adopting Open Science practices" into two separate questions: "Identify the advantages of adopting Open Science practices" and "Identify disadvantages of adopting Open Science practices." |
|  | Removal of Questions | Remove the question: "Indicate your role (integrated researcher or collaborating researcher)." |
| Adjustments to Order and Structure | Reordering Questions | Rearrange the order of some questions for better flow: The question "Are you affiliated with a research center?" should precede the question about specifying the group/center. |
|  | Conditional Advances | If the response is negative, skip to question 9. |
| Wording and Response Options Alterations | Wording Refinement | In question 13, replace: "How familiar are you with the concept of open access? (select one)? (Familiar/Not familiar)" with "Do you know what open access is? (Yes/No)." |
| Addition of Definitions and Options | Inclusion of Definitions | Add "I have never searched for information" to cover more response scenarios. |
|  | Addition of Options | In question 19, add the option "None." |

Introduction of New Questions: New questions were incorporated to address aspects such as the resources that facilitate the adoption of OS practices. Furthermore, new response options were introduced to broaden the range of possible answers, including options like “None” and “I have never searched for information.”

Division of Extensive or Complex Questions: Questions deemed lengthy or complex were segmented to enhance understanding and facilitate data analysis. For example, a question requesting the identification of both the advantages and disadvantages of OS was split into two separate questions.

Removal of Less Relevant Questions: Questions considered less relevant to the study’s objectives were removed. For example, the question about the participant’s status as an integrated or collaborating researcher was excluded.

Reorganization for Logical Flow: The questionnaire’s flow was restructured to ensure a more logical and natural sequence. For example, questions related to affiliation with research centers were repositioned for better alignment.

Rewording for Clarity and Objectivity: Some questions were rephrased to improve clarity and precision. A significant change was rewording a question about familiarity with the concept of open access into a more direct question: “Do you know what open access is? (Yes/No).”

Addition of Definitions and Examples: Definitions and examples were added to certain response options to enhance participant comprehension.

These changes were carefully planned to improve the clarity, efficiency, and alignment of the questionnaire with the research objectives, ultimately facilitating the collection of more precise and relevant data.

### Participant Characterization

The questionnaire was completed by 35 participants from the institution, out of a total population of 747 individuals (faculty and students). The analysis revealed that most respondents held faculty positions, with 33.3% being Adjunct Professors, 25.0% Coordinating Professors, 25.0% master’s Students, and 11.1% Guest Assistants, while 5.6% were students.

Regarding academic qualifications, 50.0% of participants held a Ph.D., 25.0% held a master’s degree, and 22.2% held a bachelor’s degree, while a small percentage (2.8%) did not specify their academic degree.

Concerning participation in research groups or centers, 61.1% of the participants reported being affiliated with a group or center, while 38.9% indicated no involvement in such activities. Among those affiliated, their roles as researchers were examined: 55.6% were identified as integrated researchers, while 44.4% were collaborating researchers.

When asked about research project coordination, 30.6% of participants stated they coordinated at least one project. Among these, 54.5% identified as principal investigators, while 45.5% worked as collaborating researchers within their projects. Additionally, most of the projects mentioned were national in scope (72.7%), while 27.3% were international. These findings suggest a predominance of projects focused on local contexts but with growing initiatives for large-scale collaboration.

### Open Science Practices

A key question in the questionnaire examined participants’ agreement with the principle of OS, as expressed in the statement: “Knowledge is for everyone and belongs to everyone” The results showed unanimous agreement, with 100% of respondents supporting this view. This consensus reflects a strong acceptance of OS core values within the academic community. When asked to provide reasons supporting their response, several themes emerged:

Ethics and Social Responsibility: Participants emphasized the ethical obligation to make knowledge accessible for the benefit of human and social development. Quotes included “The sharing of research benefits is an important ethical principle”; “All knowledge production is for societal development”.

Democratization and Equity in Access: Participants highlighted the need to eliminate economic and social barriers to ensure equal access to knowledge. Statements included “Access to science should not depend on economic criteria” and “There should be equal opportunities in accessing scientific knowledge”.

Promotion of Scientific Progress: Participants focused on the positive impact of knowledge sharing in advancing research, enabling collaboration, and identifying scientific gaps. Examples provided included: “The sharing of information generates knowledge and promotes new research” and “Facilitating access to knowledge allows the identification of research needs”

Personal and Professional Benefits: Open knowledge was recognized as a tool for individual and collective growth, contributing to professional and personal development. Participants noted: “Only through knowledge sharing can we grow, both professionally and personally” and “Knowledge takes no space and should not be restricted to those who produce it”.

Citizen Empowerment: Some respondents emphasized the responsibility of making research results accessible through clear and comprehensive language, promoting scientific citizenship. Statements included: “Research results should be accessible to all citizens in simple language” and “Access to credible information empowers citizens”.

Most participants reported having prior experience with at least one of the OS practices mentioned. The most frequently cited practices included open access to scientific publications (e.g., journals and repositories), open educational materials, and collaboration/replication across various locations. Conversely, practices such as pre-registration of studies, report registration, and open code were among the least experienced, suggesting potential unfamiliarity or lower adoption of these initiatives within the HEI context (Figure 1).

**Figure 1.**
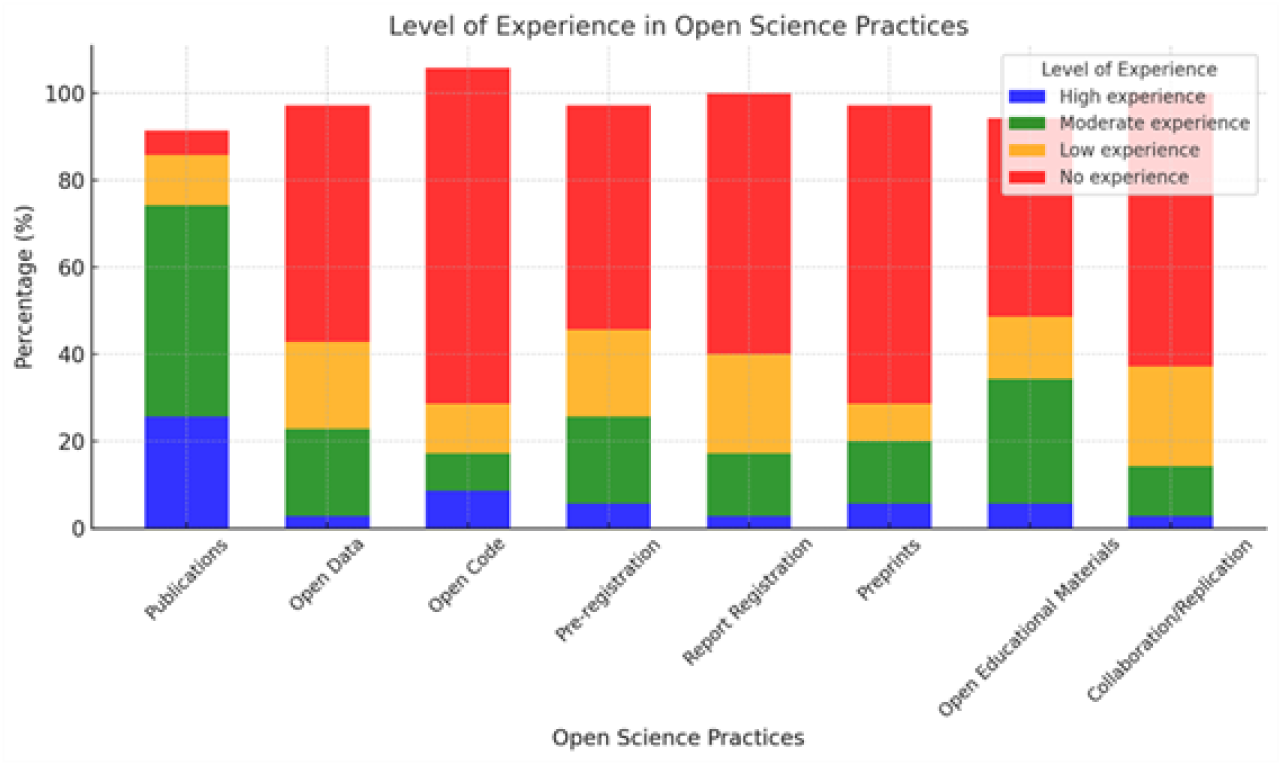
Open Science Practices.

When asked about their knowledge of open access policies, 75.0% of respondents indicated familiarity with such policies, while 25.0% stated they were unaware of them. Regarding understanding the open access concept, 63.9% of participants reported they had a good understanding, while 36.1% indicated having only a basic notion.

Most participants (28 responses, corresponding to 77.8% of the total) stated that they either use or produce open access scientific publications. In contrast, 6 participants (16.7%) expressed uncertainty about the practice, while only 2 participants (5.5%)

### Tools and Platforms

Regarding the use of databases or bibliographic research platforms, the most frequently cited platforms included the Virtual Library of the HEI (n=31), EbscoHost (n=35), Medline (n=32), Scopus (n=30), and Web of Science (n=27), demonstrating their central relevance to the participants. Approximately 30% of participants reported using the Institutional Repository, followed by the Open Science Framework (OSF, 15%). Tools such as OpenUP Hub (4.3%), Zenodo (2.1%), and others, such as ARGOS/OpenDMP (2.1%) were mentioned less frequently.

### Perceptions and Support Needs

When asked about the perceived advantages and disadvantages of adopting OS practices, participants identified several key benefits. The main advantages included increased visibility of publications and data, easier access to knowledge, and the promotion of scientific collaboration. However, participants noted significant disadvantages, including time constraints, the undervaluation of OS practices within scientific evaluation systems, and the need to adapt to new methodologies and tools. These challenges represent barriers that must be addressed to successfully implement and consolidate OS practices.

The results indicate that the greatest need for support and information on open access is related to Open Access Publishing (11.4%), followed by Applying OS Recommendations in Research Practices (9.5%) and Meeting OS Requirements During Grant Proposal Preparation (9.0%). Copyright and Open Access (7.6%) were also highlighted, underscoring the importance of these areas for the participants. These results are summarized in Figure 2.

**Figure 2.**
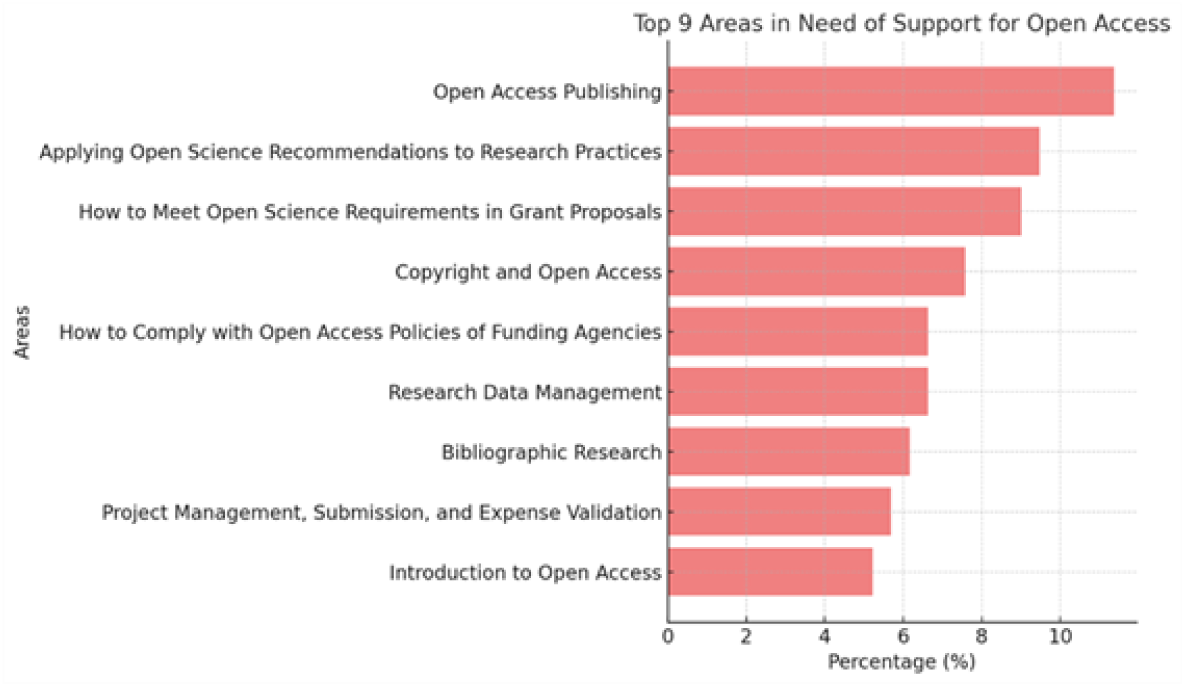
Support Needs for Open Access.

Among the most frequently mentioned support actions, the publication of educational materials in open access was the most prominent, cited by 77.14% of participants, highlighting the importance of freely sharing educational resources. Additionally, 74.29% of respondents indicated the need for technical guidance on preparing data for deposit in open repositories, reflecting the growing recognition of the importance of data transparency and accessibility.

Furthermore, 60% of participants mentioned preparing digital materials and software code for deposit in open access, demonstrating significant interest in the dissemination of technological resources. Similarly, 60% reported the need for support in the preregistration of high-quality protocols, reinforcing the importance of reproducibility and scientific integrity.

## Discussion

The Foundation for Science and Technology, I.P. (FCT), an agency under the Ministry of Education and Science of Portugal responsible for evaluating and funding scientific research activities across the country in all scientific areas, adopted the Policy on Open Access to Scientific Publications in 2014 for publications resulting from R&D Projects Funded by FCT. However, despite these advancements, such practices are not currently implemented in some contexts.

Despite the dissemination strategies implemented by the research team, adherence to the questionnaire was notably low. These findings can be attributed to the decision to administer the questionnaire via electronic mail, which, according to Hernandez et al [23] presents a disadvantage. On the other hand, this approach enables reducing bias in responses, as respondents are not influenced by the physical presence of the researcher. In preparing the questionnaire, the research team considered the need for clarity, objectivity, and conciseness in the formulation of questions, ensuring that response tendencies were not introduced and that the length of the questionnaire adhered to recommended guidelines to improve adherence rate and the quality of responses.

Another reason for the low adherence may be related to the perceived relevance and impact of OS within the institution, as members of this academic community may be little aware of the importance of OS. The evidence of low experience with OS practices, coupled with the respondents’ low level of knowledge in this area, may also contributed to deter potential respondents from engaging with the questionnaire.

Despite the participants’ agreement with the principles of OS, there remains some confusion on the topic, leading to paradoxical results. For example, although most of the respondents reported knowing open access principles and having utilized them or having contributed to open access publications, many also indicated a lack of knowledge about open access policies. Moreover, not all participants were familiar with the various open access formats and policies. These findings suggest that although there is considerable alignment with the concept of OS, gaps in knowledge and information persist. Addressing these gaps through targeted training initiatives and awareness campaigns could help bridge these gaps. Although no similar studies were found involving Nursing HEI, previous research has reported mixed findings on researchers’ perceptions and practices, according to different contexts [24,25].

These study findings underscore the need for investment in skills training addressing OS practices and for in-depth studies on motivation, adherence and the conditions for the development of OS practices, particularly within the context of higher education, in the health area. These results are consistent with those reported in similar studies [24].

In view of the above stated, it is essential to design training programs that allow immersion in OS environments, thus promoting the development of critical skills, namely the ability to evaluate and analyze information sources, encouraging a proactive approach to the construction of knowledge. Active participation in scientific production, through collaboration in accessible and open research projects, and promotes immersion in the process of discovery and innovation, empowers constructive thinking and problem-solving skills that transcend disciplinary boundaries. Ultimately, such initiatives contribute to meaningful and transformative learning experiences [26–30].

The study provided valuable contributions to the institution in which it was conducted, not only as a catalyst for advancing OS practices but also as a means of fostering awareness and highlighting the need for investment and support in establishing a robust research framework conducive to further investigation and improvement of OS practices. Additionally, this study serves as a benchmarking reference for similar studies in other higher education contexts, including the health field, where concerns regarding privacy and confidentiality present significant barriers to implementation among others.

This study was limited by the low participation rate in the questionnaire, hindering the generalizability of the results to the broader population under study. However, despite this limitation, this study has identified valuable opportunities and provided evidence that can inform future research aimed at addressing these constraints. More importantly, these study findings serve as a crucial foundation for guiding the implementation of OS practices and the development of further studies in this field.

## Conclusion

This study offers valuable insights into the adoption of OS practices within a higher education institution. While open access publishing is widely accepted, other key practices, such as study preregistration and open-source coding remain less familiar. Furthermore, participants predominantly rely on traditional academic databases, with OS-specific platforms being underutilized.

These findings highlight the need for targeted training initiatives, particularly in open access publishing, data management, and the implementation of OS recommendations.

Enhancing institutional support and raising awareness could facilitate a more comprehensive and effective integration of OS practices, ultimately enhancing transparency, fostering innovation, and maximizing the societal impact of scientific research.

## Data Availability

Data cannot be shared publicly because of reasons of anonymity and confidentiality. Data are available from the Nursing School of Porto / Ethics Committee (contact via) for researchers who meet the criteria for access to confidential data.

https://forms.office.com/e/qrGdPRkX36?origin=lprLink

## References

1. European Commission. Open innovation, open science, open to the world: a vision for Europe [Internet]. Brussels: European Commission; 2016. Available from: https://publications.europa.eu/en/publication-detail/-/publication/3213b335-1cbc-11e6-ba9a-01aa75ed71a1. DOI: 10.2777/061652. Consulted at: November 28, 2023.

2. Chavanayarn S. The epistemic value of open science. Open Sci J. 2018;3(3).

3. ERC Scientific Council. Open Research Data and Data Management Plans: Information for ERC Grantees. Version 4.1, April 20, 2022 [Internet]. Available from: https://erc.europa.eu/managing-your-project/open-science. Accessed on: January 18, 2025.

4. Rahman NA. The Need for Open Science. J Res Manag Gov. 2019;2(1):22–30.

5. Fouilloux A, Trasatti E, Foglini F, Coca-Castro A, Iaquinta J. FAIR research objects for realising open science with the EOSC project RELIANCE. Res Ideas Outcomes. 2023.

6. Hofmann B. Open Science knowledge production: Addressing epistemological challenges and ethical implications. Publ. 2022;10(3):24.

7. De Rosa R, Aragona B. Open science and the academic profession. JeDEM - eJournal of eDemocracy Open Gov. 2021;13(2).

8. Vicente-Saez R, Gustafsson R, Van den Brande L. The dawn of an open exploration era: Emergent principles and practices of open science and innovation of university research teams in a digital world. Technol Forecast Soc Change. 2020;156:120037.

9. Blanco A. The role of Open Science in our research. BioResources. 2024;19(2):2013–6.

10. Maciuliene M. Beyond Open Access: Conceptualizing Open Science for knowledge co-creation. Front Commun. 2022;7.

11. Cole NL, Kormann E, Klebel T, Apartis S, Ross-Hellauer T. The societal impact of Open Science: A scoping review. R Soc Open Sci. 2024;117.

12. Silva, R. C. G. D., Cardoso, D. F. B., Cardoso, M. L. D. S., Sá, M. D. C. G. M. A. D., & Apóstolo, J. L. A. Citizen involvement in scientific activities and extension of knowledge to society. Revista da Escola de Enfermagem da USP. 2021; 55, e20210171.

13. Abele-Brehm AE, Gollwitzer M, Steinberg U, Schönbrodt FD. Attitudes toward open science and public data sharing. Soc Psychol. 2019;50.

14. Sullivan I, DeHaven A, Mellor D. Open and reproducible research on Open Science Framework. Curr Protoc Essent Lab Tech. 2019;18.

15. Van Dijk, W., Schatschneider, C., & Hart, S. A.. Open Science in Education Sciences. Journal of Learning Disabilities. 2021; 54(2), 139–152.

16. Stojanovski, J., Aspaas, P. P. Open Science – A Croatian Perspective. Open Science Talk. 2022; No. 45.

17. OECD. Making Open Science a Reality. Organisation for Economic Co-operation and Development. 2015 [Internet]. Available from: https://www.oecd.org/content/dam/oecd/en/publications/reports/2015/10/making-open-science-a-reality_g17a270f/5jrs2f963zs1-en.pdf Accessed on: January 18, 2025.

18. United Nations Educational, Scientific and Cultural Organization (UNESCO). UNESCO Recommendation on Open Science. UNESCO Publishing. 2022 [Internet] Available from: https://unesdoc.unesco.org/ark:/48223/pf0000379949. Accessed on: January 18, 2025.

19. Saraite-Sariene, L., Caba-Pérez, C., & López-Hernández, A. M. Expanding the actions of Open Government in higher education sector: From web transparency to Open Science. PLOS ONE. 2020; 15(9), e0238801. 10.1371/journal.pone.0238801.

20. Von Elm, E., Altman, D. G., Egger, M., Pocock, S. J., Gøtzsche, P. C., & Vandenbroucke, J. P. The Strengthening the Reporting of Observational Studies in Epidemiology (STROBE) statement: guidelines for reporting observational studies. The lancet. 2007; 370(9596), 1453–1457.

21. Marques CT, Barros C, Camacho M, Contreiras P. Acesso Aberto: conhecimento e práticas na NOVA FCSH. Relatório de análise ao inquérito por questionário. Lisboa: Faculdade de Ciências Sociais e Humanas, Universidade Nova de Lisboa; 2021.

22. Science Europe Working Group on Open Access. Science Europe Principles on Open Access to Research Publications [Internet]. Belgium; 2015. Available

23. Hernández Sampieri R, Fernández Collado C, Baptista Lucio P. Metodología de la Investigación. México: McGraw-Hill; 2000.

24. Ollé C, López-Borrull A, Melero R, Boté-Vericad JJ, Rodríguez-Gairín JM, Abadal E. Habits and perceptions regarding open science by researchers from Spanish institutions. PLoS ONE. 2023;18(7):e0288313. 10.1371/journal.pone.0288313

25. Pownall M, Terry J, Collins E, Sladekova M, Jones A. UK Psychology PhD researchers’ knowledge, perceptions, and experiences of open science. Cogent Psychol. 2023;10(1). 10.1080/23311908.2023.2248765

26. Antunes MDL, Lopes C, Sanches T. Literacia da informação e ciência aberta em saúde: o antes e o depois. XIII Jorn APDIS. 2018;1–16.

27. Lopes CA, Antunes MDL, Sanches T. Contributos da literacia da informação para a Ciência Aberta. Ibersid. 2018;12(1):59–67.

28. Lopes C, Antunes MDL, Sanches T. Integração das competências de literacia da informação no currículo académico: aplicação da Framework da ACRL. 2018.

29. Okada A, Rodrigues E. A educação aberta com ciência aberta e escolarização aberta para pesquisa e inovação responsáveis. Educ Fora Caixa: Tend Intern Perspect Sobre Inov Educ. 2018; 4:41–54.

30. Rocha D, Loureiro A. Ciência Aberta e Ciência Cidadã no IPSantarém: responsabilidade social na investigação. Encontro Nac Ciên Cidadã. 2021. from: https://www.scienceeurope.org/media/4kxhtct2/se_poa_pos_statement_web_final_20150617.pdf

